# Lessons learned from implementation of interferon-gamma release assay to screen for latent tuberculosis infection in a large multicenter observational cohort study in Brazil

**DOI:** 10.1101/2021.08.10.21261605

**Authors:** Allyson G. Costa, Brenda K.S. Carvalho, Mariana Araújo-Pereira, Hiochelson N.S. Ibiapina, Renata Spener-Gomes, Alexandra B. Souza, Adriano Gomes-Silva, Alice M.S. Andrade, Elisangela C. Silva, María B. Arriaga, Aline Benjamin, Michael S. Rocha, Adriana S.R. Moreira, Jamile G. Oliveira, Marina C. Figueiredo, Megan M. Turner, Betina Durovni, Solange Cavalcante, Afranio L. Kritski, Valeria C. Rolla, Timothy R. Sterling, Bruno B. Andrade, Marcelo Cordeiro-Santos, the RePORT Brazil Consortium

## Abstract

**Background:** Interferon-gamma release assay (IGRA) has emerged as a useful tool in identifying latent tuberculosis infection (LTBI). This assay can be performed through testing platforms, such as QuantiFERON-TB Gold Plus (QFT^®^-Plus). This in vitro test has been incorporated by several guidelines worldwide and has recently been considered for the diagnosis of LTBI by the World Health Organization (WHO). The possibility of systematically implementing IGRAs such as QFT^®^-Plus in centers that perform LTBI screening has been accelerated by the decreased availability of tuberculin skin testing (TST) in several countries. Nevertheless, the process to implement IGRA testing in routine clinical care has many gaps.

**Methods:** The study utilized the expertise acquired by the laboratory teams of the Regional Prospective Observational Research in Tuberculosis (RePORT)-Brazil consortium during study protocol implementation of LTBI screening of TB close contacts.

**Results:** RePORT-Brazil includes clinical research sites from Brazilian cities and is the largest multicenter cohort of TB close contacts to date in the country. Operational and logistical challenges faced during IGRA implementation in all four study laboratories are described, as well as the solutions that were developed and led to the successful establishment of IGRA testing in RePORT-Brazil.

**Conclusions:** The problems identified and resolved in this study can assist laboratories implementing IGRAs, in addition to manufacturers of IGRAs providing effective technical support. This will facilitate the implementation of IGRA testing in countries with a high TB burden, such as Brazil.

**Importance:** The interferon-gamma release assay (IGRA) has emerged as a useful tool in identifying persons with latent tuberculosis infection (LTBI). Although the implementation of IGRAs is of utmost importance, to our knowledge, there is scarce information on identification of logistical and technical challenges of systematic screening of for LTBI on a large scale. Thus, the problems identified and resolved in this study can assist laboratories implementing IGRAs, in addition to manufacturers of IGRAs providing effective technical support. This will facilitate the implementation of IGRA testing in countries with a high TB burden, such as Brazil.

## 1 Introduction

The World Health Organization (WHO) estimates that one quarter of the global population is infected with *M. tuberculosis* (Mtb) (1, 2). Most individuals exposed to Mtb who become infected are asymptomatic, referred to as latent tuberculosis infection (LTBI). Between 5 to 10% of individuals with LTBI, if not treated with tuberculosis preventive therapy (TPT), can progress to active tuberculosis (TB) during their lifetime (3, 4). Thus, diagnosis and treatment of LTBI is critical to reduce the incidence of active TB and control Mtb transmission.

In several countries endemic for TB, such as Brazil, screening for LTBI has traditionally utilized the tuberculin skin test (TST), which consists in the intradermal inoculation of the TB-purified protein derivative (PPD) and evaluation for cutaneous induration. However, this test has several limitations, such as false-positive reactions in persons with BCG vaccination or infection with non-tuberculosis mycobacteria, and false-negative reactions in persons with immune suppression (5, 6). Additional issues include the requirement for a return visit to assess the skin for induration within 48-72 hours after PPD inoculation, and the subjective interpretation of the dermal reaction (i.e., inter-reader variability) (7, 8). Recent scientific advances in molecular investigations allowed the isolation of Mtb-specific antigens that drive production of interferon-gamma (IFN-γ) by specific T-lymphocytes, enabling the development of more specific assays based on cellular recall responses, to identify LTBI (9, 10).

The interferon-gamma release assay (IGRA) has emerged as a useful tool in identifying persons with LTBI. This assay can be performed through two distinct testing platforms, such as the enzyme-linked immunosorbent spot (ELISPOT) assay (T-SPOT.TB) or QuantiFERON-TB Gold Plus (QFT^®^-Plus). These in vitro tests have been incorporated into several guidelines worldwide and have recently been considered equivalent for the diagnosis of LTBI by the WHO (9, 10). Of note, both IGRA tests have advantages over TST in several aspects, including: (i) they do not require a follow-up visit for obtaining results, and (ii) they use in vitro stimulation of cells from the peripheral blood with Mtb-derived ESAT-6 and CFP-10 proteins, which are absent in the BCG vaccine as well as in most non-TB mycobacteria, resulting in a higher specificity (11–13). The advantages of the IGRA over the TST indicate that this immunoassay may be a reliable alternative. On the other hand, the use of IGRA results on high costs, the necessity to be carried out in sites using good clinical and laboratory practices and well-trained technical personnel and available equipment’s.

The advantages of the IGRA over the TST indicate that this immunoassay may be a reliable alternative to TST. The possibility of systematically implementing IGRAs such as QFT^®^-Plus in centers that perform LTBI screening has been accelerated by the decreased availability of TST in several countries, including Brazil (14). Implementation of QFT^®^-Plus in TB reference centers could facilitate screening, diagnosis, and treatment of LTBI, and thereby reduce the TB burden.

Although the implementation of IGRAs is of utmost importance, to our knowledge, there is scarce information on identification of logistical and technical challenges of systematic screening of for LTBI on a large scale. The present study was designed to fill this gap and provide information that would improve the effectiveness and efficiency of IGRA-based LTBI screening. The operational and logistical challenges faced during IGRA implementation in all four study laboratories are also described, as well as the solutions that were developed and which led to the successful establishment of IGRA testing in RePORT-Brazil.

## 2 Material and Methods

### 2.1 Study Design and Laboratory Sites

The present investigation was an implementation study performed within a multicenter cohort study (RePORT-Brazil) between 2016 and 2019. It was conducted in five research centers located in four Brazilian cities: Fundação de Medicina Tropical Dr. Heitor Vieira Dourado (FMT-HVD) in Manaus-Amazonas, Instituto Brasileiro para Investigação da Tuberculose (IBIT) in Salvador-Bahia, Secretaria Municipal de Saúde de Duque de Caxias (SMS-DC) in Duque de Caxias-Rio de Janeiro, Instituto Nacional de Infectologia Evandro Chagas (INI) and Clínica da Família Rinaldo Delamare, Secretaria Municipal de Saúde do Rio de Janeiro (SMS-RJ) both in Rio de Janeiro-Rio de janeiro (15). The five health centers are located in 3 distinct regions of Brazil, with similar climate conditions (equatorial and tropical), temperature and humidity, showing range 19.9°C to 26.4°C and 77.2 to 85.1%, respectively. (16, 17).

### 2.2 Maintenance and Biosafety of Laboratory Sites

Standard Operating Procedures (SOPs), Good Clinical Practice (GCP), Good Clinical Laboratory Practices (GCLP) and other trainings were carried out by the project staff prior to initiating the study. In addition, all sites provided up to date equipment maintenance certifications to ensure test quality and to minimize risks for laboratory technicians who processed biologicals daily.

### 2.3 QuantiFERON-TB Gold Plus (QFT®-Plus)

Initially QFT-TB Gold in Tube (QFT-GIT) was implemented into the laboratory routine of the RePORT-Brazil consortium, subsequently replaced by QFT^®^-Plus. Every laboratory received training on sample collection and processing by the QIAGEN corporation (Chatsworth, CA, USA). Supplementary Figure 1 summarizes QFT^®^-Plus steps. Briefly, venous blood for testing was collected in four tubes (Nil, TB1, TB2 and Mitogen) and incubated at 37°C for 20h. After incubation, samples were stored at -20°C until the enzyme-linked immunosorbent assay (ELISA) was performed within 2 weeks. IFN-γ levels (international units per milliliter [IU/mL]) were quantified with a 4-point standard curve. QFT^®^-Plus Analysis Software was used to generate the results according to the manufacturer’s recommendations (18). The software performed a quality control assessment of the assay, generated a standard curve, and provided quantitative (IU/mL) and qualitative (positive, negative, or indeterminate) results.

### 2.4 Laboratory data of the QFT^®^-Plus implementation process

Laboratory information from the QFT^®^-Plus implementation process was evaluated by the teams from all RePORT-Brazil laboratories. Data were obtained from team training, equipment maintenance, and pre-analytical evaluations, such as: type of sample collection, place of sample collection and processing, tube identification (ID), transport quality control (types of transport, time and temperature of the samples), and presence of hemolysis, clots and volume of the samples, as well as qualitative results (number of positive, negative and indeterminate tests). All data were entered into the Research Electronic Data Capture (REDCap) platform, reviewed by data managers for quality control (QC), and subsequently approved for the study analyses.

### 2.5 Descriptive and Statistical Analysis

Descriptive analyses were performed to characterize the study labs and quality control measurements. Categorical variables were displayed as frequency and percentages and compared using a two-sided Pearson’s chi-square test (Yate’s correction) or the Fisher’s two-tailed test in 2×3 or 2×2 tables, respectively. Continuous variables were displayed as median and interquartile ranges (IQR) and tested for Gaussian distribution using the D’Agostino-Pearson test. Comparisons of values between two groups of data were performed using the Mann-Whitney U test. The Spearman rank correlation test was carried out to assess relationships between variables. Data analyses were performed using R (version 3.6.3), using Hmisc (version 4.4.1), compare Groups (version 4.4.3), ggplot2 (version 3.3.2) and ggcorrplot (version 0.1.3) R packages. All analyses were prespecified. Differences with p-value <0.05 were considered statistically significant.

### 2.6 Ethical Approval

The study was conducted according to the principles of the Declaration of Helsinki. The RePORT-Brazil protocol was approved by the institutional review boards at each study site and at Vanderbilt University Medical Center. In additional, the present study was submitted to and approved by the Ethical Committee at Instituto de Pesquisa Clínica Evandro Chagas, Fundação Oswaldo Cruz (FIOCRUZ), under the protocol registration number #688.067, Secretaria Municipal de Saude do Rio de Janeiro (SMS/RJ), under the protocol registration number #740.554, Hospital Universitario Clementino Fraga Filho, under the protocol registration number #852.519, Maternidade Climério de Oliveira (MCO), Universidade Federal do Bahia (UFBA), under the protocol registration number #723.168 and Fundação de Medicina Tropical Dr. Heitor Vieira Dourado (FMT-HVD), under the protocol registration number #807.595. The patients, parents or legal guardians read and signed the informed consent form prior to inclusion of the patients in the study. The study fulfills the principles of the Helsinki declaration and the 466/2012 resolution of the Brazilian National Health Council for research involving human participants.

## 3 Results

### 3.1 Interferon-gamma release assay implementation

The IGRA/QFT^®^-Plus implementation process started with the checklist provided by QIAGEN Corp. The four laboratories proceeded with the acquisition of equipment (micropipettes, 37°C incubator, centrifuge, refrigerator, freezer -20°C, microplate washer, microplate reader and computer), reagents (ultrapure dH2O and cleaning solutions), consumables (tips, microtubes and solution reservoir), QFT^®^-Plus Tube kit and QFT^®^-Plus ELISA kit. Subsequently, laboratory technicians took GCLP and were trained on the steps of collecting, transporting, incubating, and processing samples for laboratory tests. In addition, QFT^®^-Plus ELISA kit assays were performed, and a pilot test was carried out, with the objective of including improvement strategies to initiate the study protocol (**Figure 1**).

**Figure 1:**
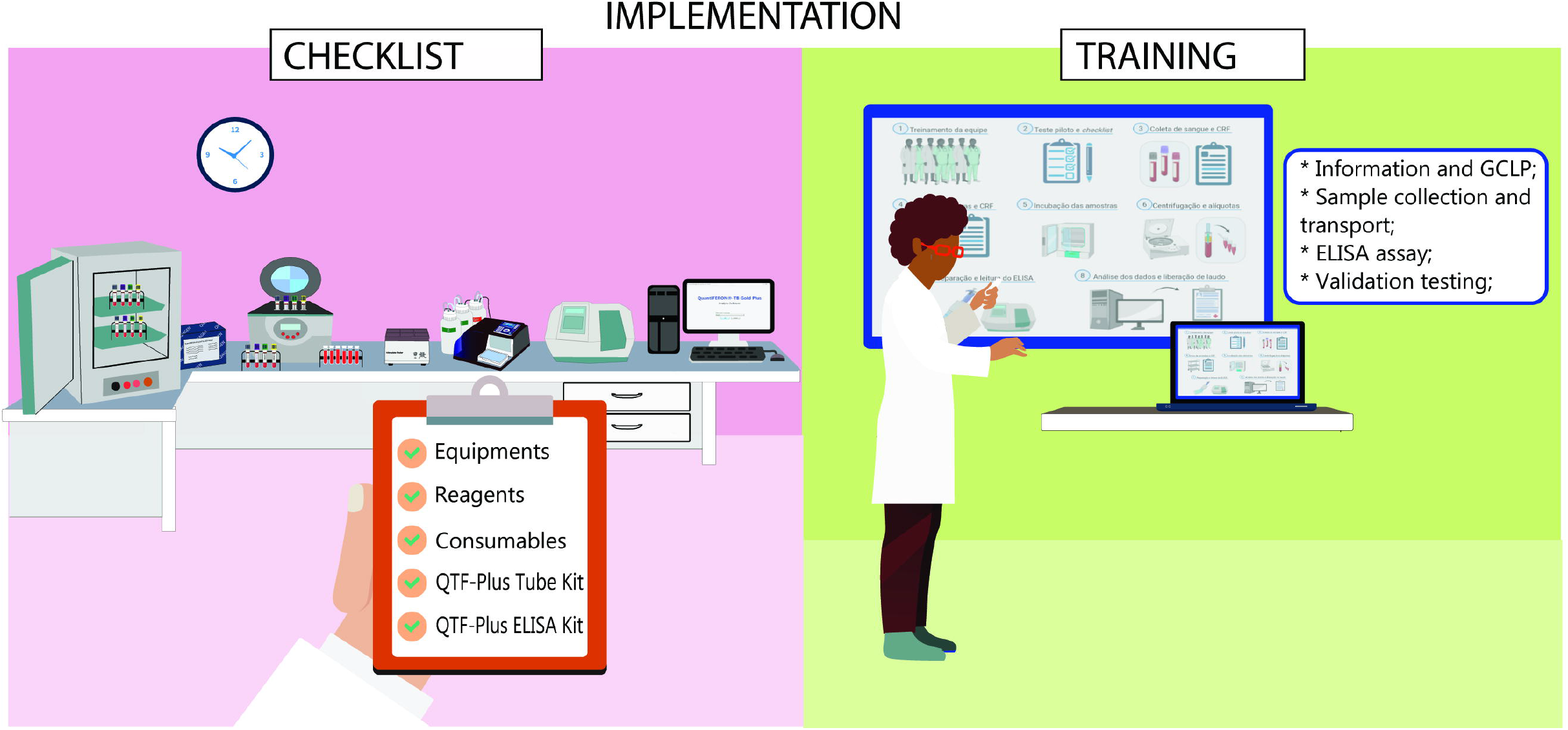
Description of implementation of IGRA/QFT^®^-Plus in the study. The first step was to check if all equipment and reagents were available and in good condition. Then, the team was trained to perform the test following Good Clinical Laboratory Practices and according to the manufacturer’s recommendations.

During the IGRA implementation process, several gaps were identified, even after following all of the recommendations of the manufacturer of the QFT^®^-Plus assay. Initially, the laboratories identified problems related to acquisition of equipment, reagents, and consumables. The equipment had to be purchased through the sites, generating additional costs. Furthermore, the microplate washer had problems at the beginning of the ELISA assays, requiring corrective maintenance. QFT^®^-Plus Tube kits and QFT^®^-Plus ELISA kits were imported, which generated logistical problems regarding delivery to the sites. To resolve this problem, delivery of the QFT^®^-Plus Tubes and ELISA kits were directed to one of the clinical sites in Rio de Janeiro, which then distributed them to the other laboratories.

### 3.2 Interferon-gamma release assay under routine conditions

The five clinical sites started recruiting patients and collecting samples under routine conditions. Prior to starting, the collecting station was organized, accounting for the environment and the tubes to be used in the QFT^®^-Plus test. These tubes had to be stored between 4°C and 25°C and taken out for immediate use only. In addition, the tubes had to be collected in a specific order, following the manufacturer’s recommendations, with the Nil tube being collected first, followed by the TB1, TB2 and Mitogen tubes. The tubes containing the samples went through a homogenization process that consisted of several inversions, where all the biological material had to come into contact with the inner surface of the tube (L-motion with exactly 10 inversions). Finally, QFT^®^-Plus Tubes was packed in boxes with temperatures ranging from 17°C to 27°C for transfer to the processing laboratory (**Figure 2**).

**Figure 2:**
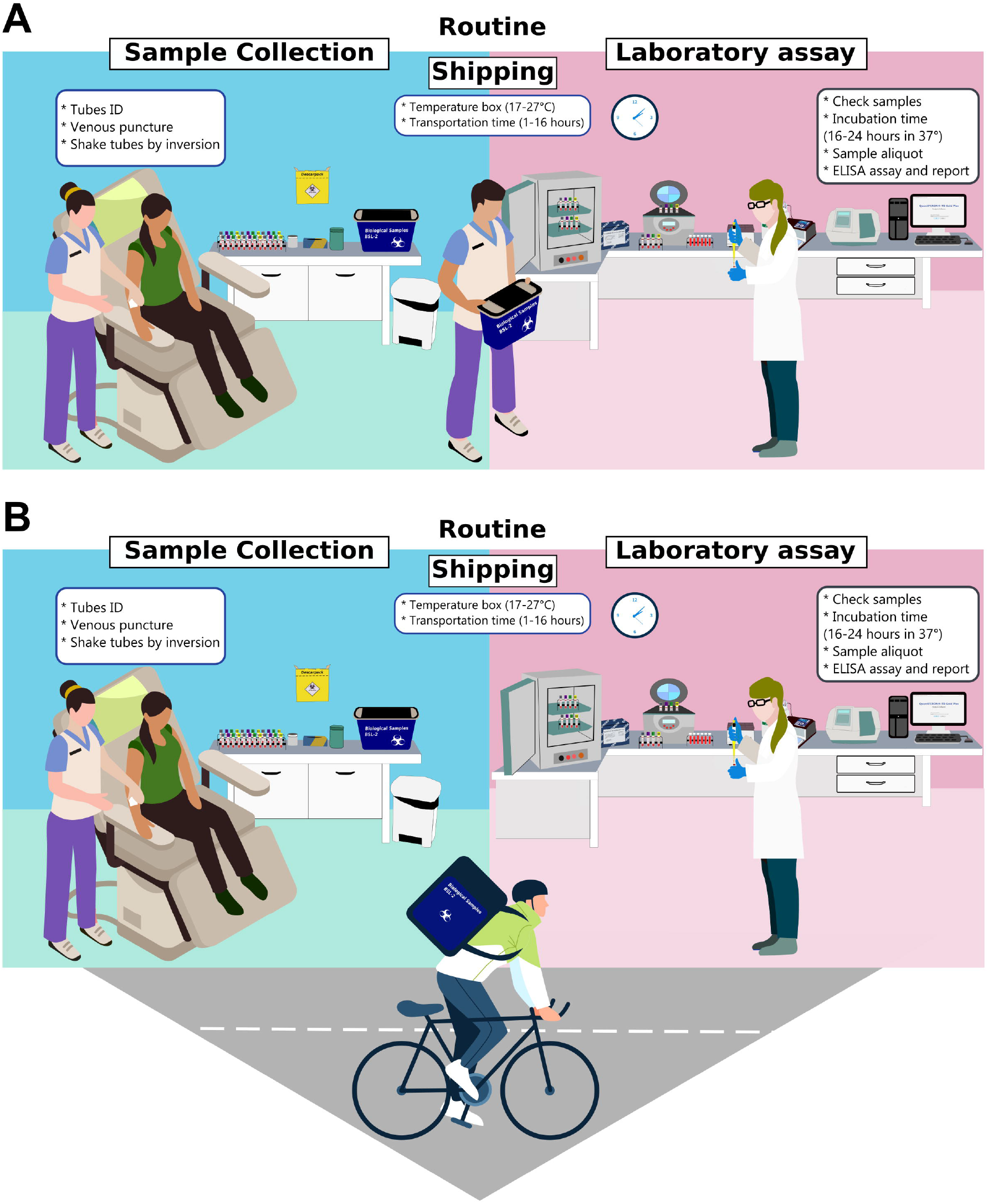
Two different conditions for IGRA/QFT^®^-Plus sample collection and processing in the study. A) Setup A: utilized by Sites 1 and 2 and characterized by performing the collection and processing of samples in the same place, without a vehicle. B) Setup B: utilized by Sites 3, 4 and 5, characterized by performing the collection and processing of samples in distinct places, and transportation of the samples by vehicle.

After collection at the clinical sites, samples were transported to the four RePORT-Brazil laboratories in two different settings, clustered in: Setup A: pertaining to sites 1 and 2, characterized by performing the collection and processing of samples in the same place, with the collection room and laboratory within the same site. Thus, the study staff responsible for collecting and organizing the samples could transport the samples to the laboratory, without requiring a vehicle (bicycle, motorcycle or car) (**Figure 2A**); Setup B: pertaining to sites 3, 4 and 5, characterized by performing the collection and processing of samples in distinct places, with the need for transportation to the laboratory and thus greater demand for time, organization and attention so that there was no excessive tube vibration. This setup required a vehicle (bicycle, motorcycle or car) to transport samples (**Figure 2B**).

Upon arrival at the labs, samples had to pass a quality check regarding transport time, temperature, tube ID, volume evaluation and the presence of hemolysis or clots. Finally, the samples were processed, incubated at 37°C for 20 hours, aliquots prepared, and the plasma samples frozen at -20°C until the QFT^®^-Plus ELISA was performed.

### 3.3 Interferon-gamma release assay results under routine conditions in two settings

**Figure 3** summarizes the IGRA/QFT^®^-Plus results under routine conditions in Setup A-B (Figure 3A) and sites 1-5 (Figure 3B) during the four-year study period (2016-2019). These results were obtained after the QFT^®^-Plus ELISA assays were performed by the teams at the sites. For this, the aliquots containing the samples were thawed and used only once. In addition to the recommendations indicated by the manufacturer, the laboratories underwent quality control with the results validated by an external laboratory. Setup B had a higher percentage of indeterminate results, mainly in the first years of implementation of QFT^®^-Plus. In addition, the number of samples changed over time for each setup, as follows: 2016 (Setup A = 190; Setup B = 173), 2017 (Setup A = 366; Setup B = 258), 2018 (Setup A = 505; Setup B = 471), 2019 (Setup A = 570; Setup B = 333). Also note that over the years of implementation, the indeterminate percentage tended to decrease. This could be associated with the learning process of site personnel. In addition, higher undetermined results were observed in Setup A in 2018 and 2019, specifically at site 1, which could be attributed to a greater number of tests performed in this setup and site.

### 3.4 Non-conformities in Interferon-gamma release assay testing

During the QFT^®^-Plus implementation process, several problems were detected, generating non-conformities, such as samples without an ID, transport with temperature outside the established standard (temperature deviation), sample leakage, and other issues (e.g. transport box change, coagulated samples, or a non-standard set of transport conditions). The proportion of non-conformities is shown in **Figure 4**.

**Figure 3:**
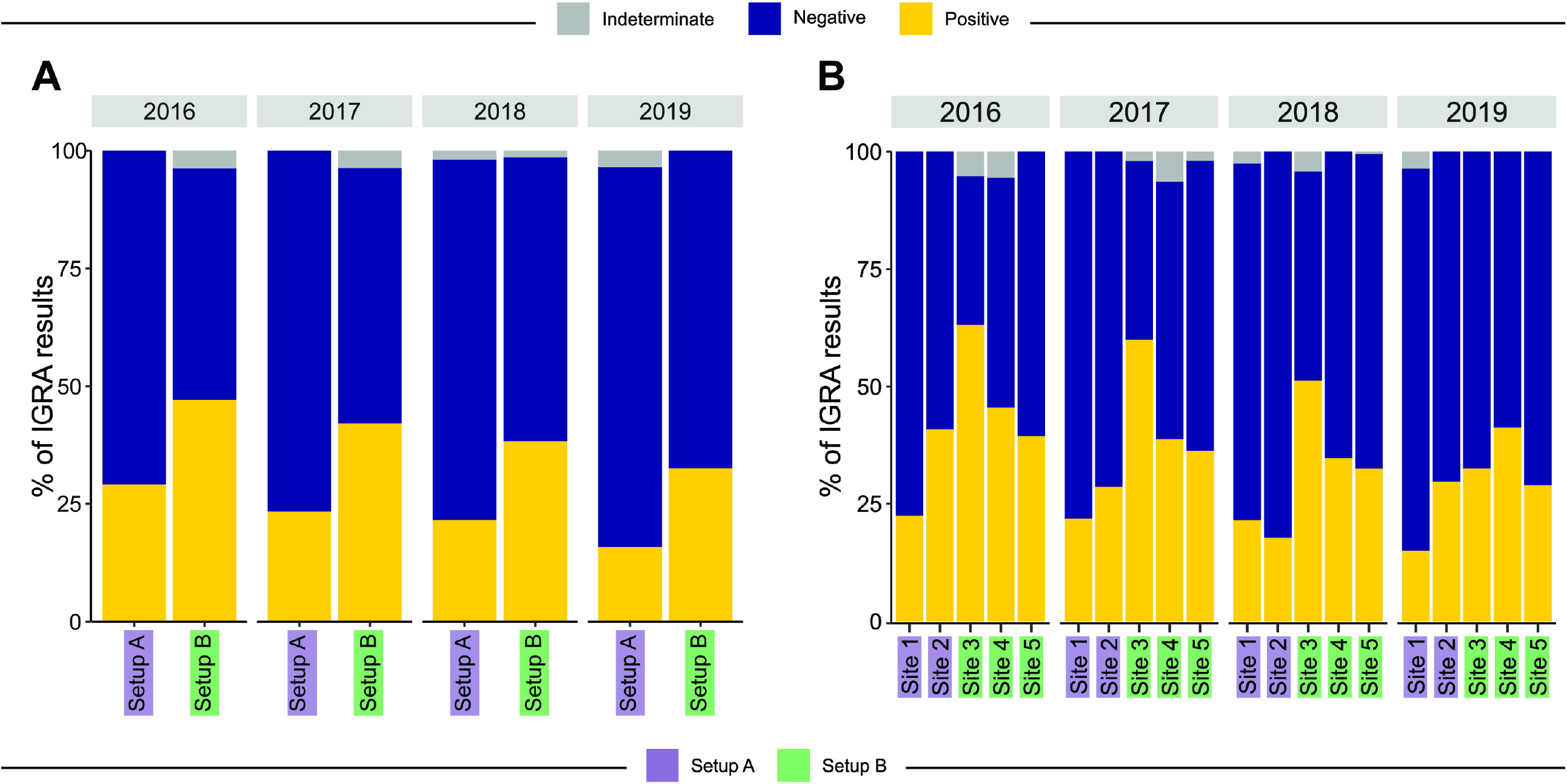
Frequencies of IGRA/QFT^®^-Plus results in each Setup and Site stratified by year of the study. A) The sites are grouped by Setup A (purple rectangles, site 1 and 2) and Setup B (green rectangles, site 3, 4 and 5). B) Results stratified by site.

**Figure 4:**
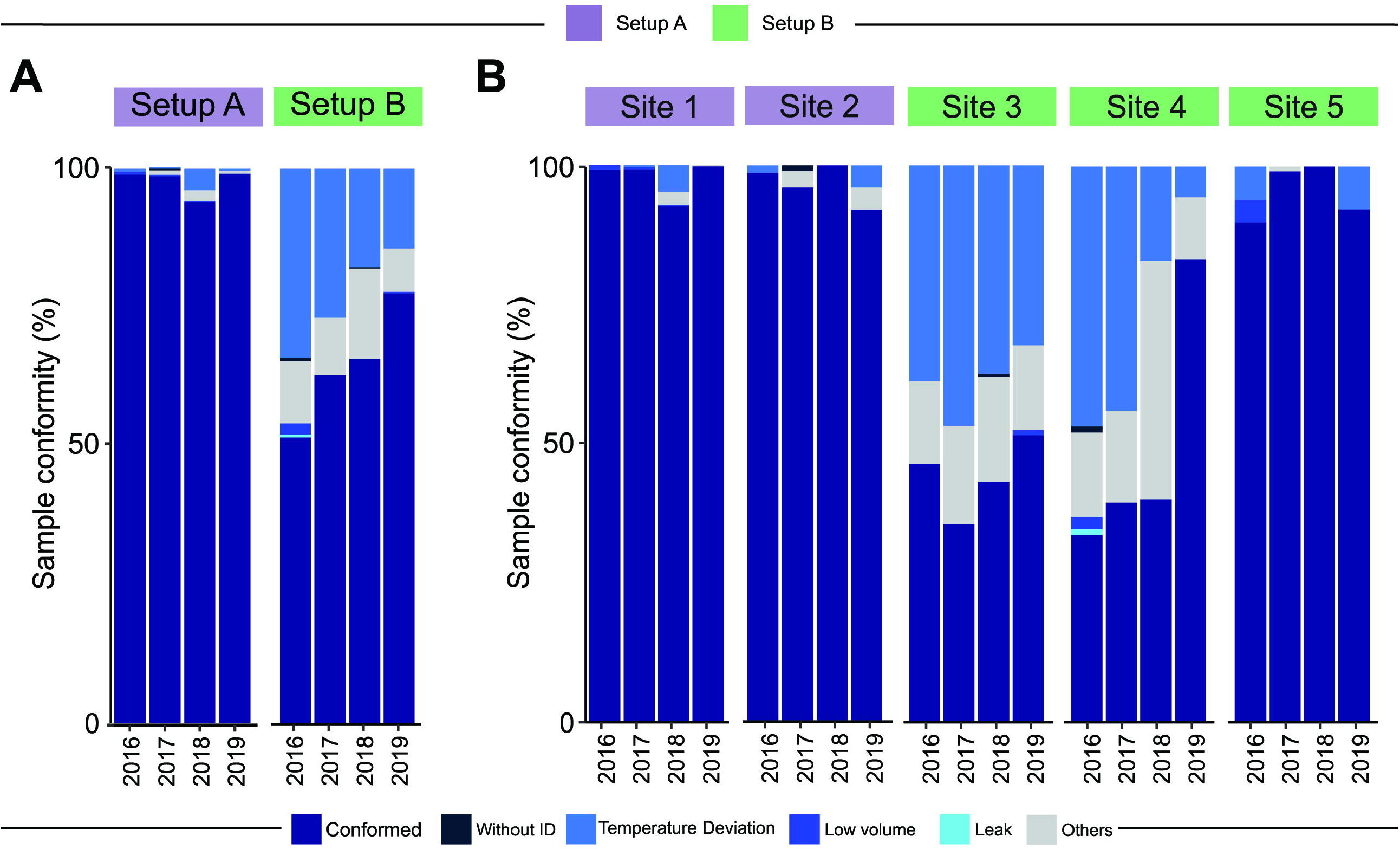
Frequencies of conformities and non-conformities of samples in each Setup and Site stratified by year of the study. A) The sites are grouped by Setup A (purple rectangles, site 1 and 2) and Setup B (green rectangles, site 3, 4 and 5). B) Results stratified by site.

The temperature deviation and other non-conformities were noted particularly among Setup B sites (**Supplementary Table 1**) compared to Setup A in all analyzed years (p<0.001). These results were likely due to conditions related to the collection and transportation of samples, since Setup B required sample transport by vehicle. In addition, these gaps generated learning opportunities for the teams, which over the years of implementation, decreased the proportion of reported non-conformities. Of note, Setup B also presented significantly higher occurrences of transport box change, coagulated samples, and lack of minimum transport conditions, characterized in the table as “Others” (p<0.001, **Supplementary Table 1**).

### 3.5 Temperature deviation was the main non-conformity in Interferon-gamma release assays

Temperature deviation was the main non-conformity identified in our study. When the occurrence was observed over the quarters of the evaluated years, it occurred mainly in the months with the highest temperatures at the sites, such as spring and summer (3^rd^, 4^th^ and 1^st^ quarter; **Figure 5**). There was a decrease in this non-conformity over the years of the study period.

**Figure 5:**
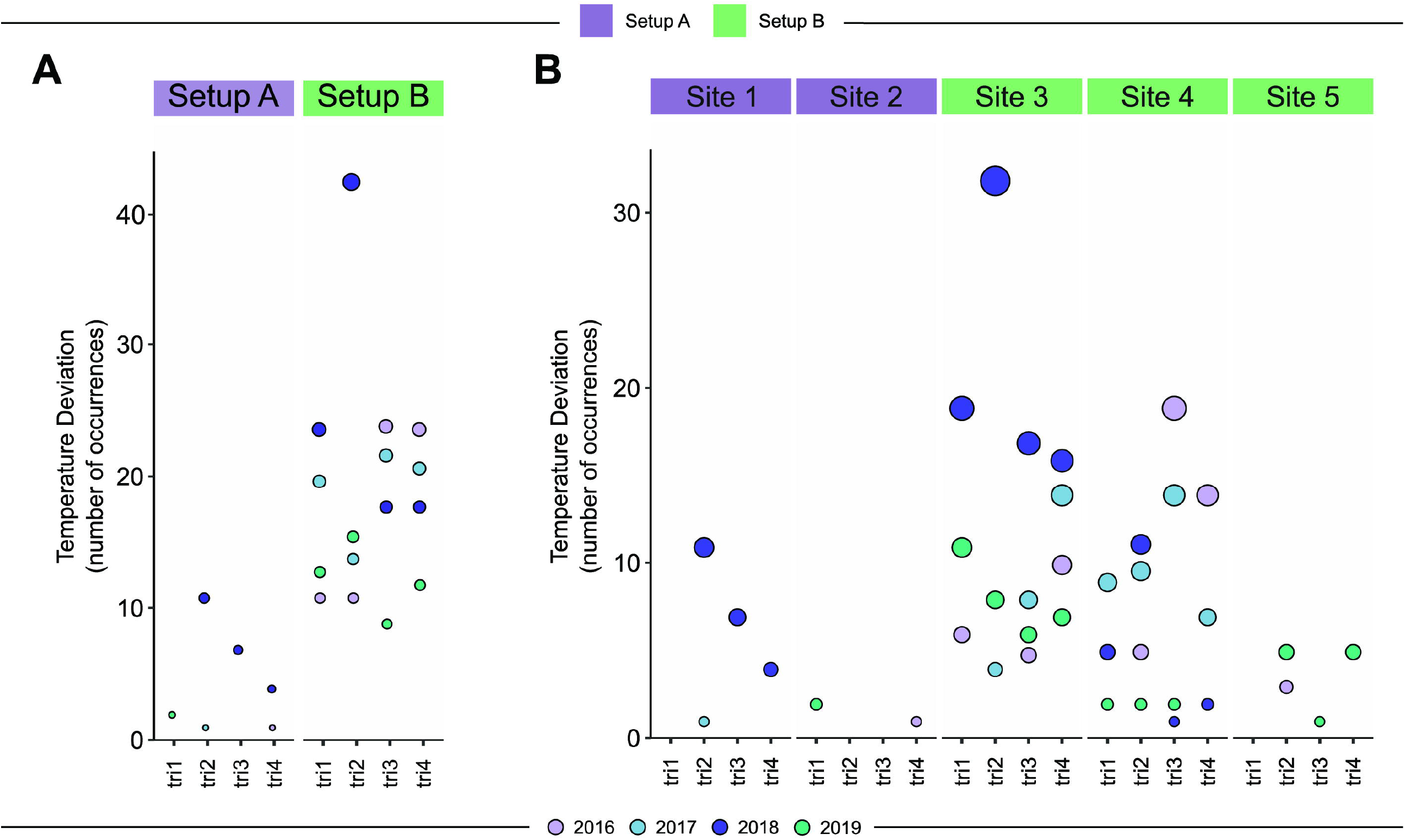
Number of occurrences of the non-conformity “Temperature deviation” in each Setup and Site stratified by year in period of study. A) The sites are grouped by Setup A (purple rectangles, site 1 and 2) and Setup B (green rectangles, site 3, 4 and 5). B) Results stratified by site. Color of circles indicates the year of non-conformity register and the size is proportional to the number of occurrences.

### 3.6 Dynamics of the time and temperature between collection and processing of Interferon-gamma release assay samples

The distance between the collection site and the sample processing laboratory directly influenced the time and the temperature variation during transportation (**Table 1**). Setup A showed a significantly shorter time between sending and receiving the samples in all studied years (p<0.001), influencing the temperature variation (delta), which was also significantly less in comparison to Setup B (p<0.001).

**Table 1:**
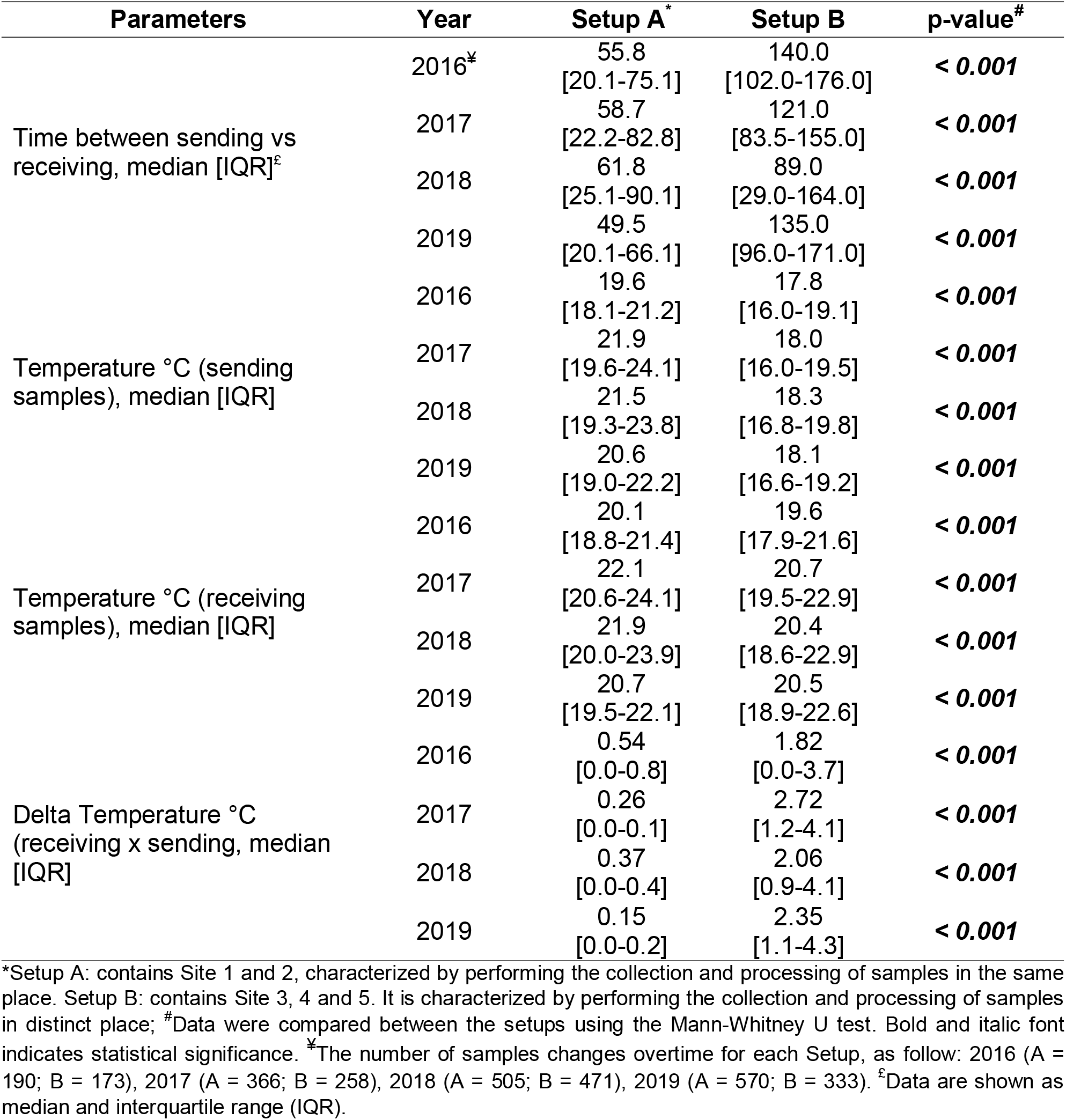
Time and temperature quality control measurements by year in period of study.

| Parameters | Year | Setup A* | Setup B | p-value<sup>#</sup> |
| --- | --- | --- | --- | --- |
| Time between sending vs receiving, median [IQR]<sup>£</sup> | 2016<sup>*</sup> | 55.8<br>[20.1-75.1] | 140.0<br>[102.0-176.0] | **< 0.001** |
|  | 2017 | 58.7<br>[22.2-82.8] | 121.0<br>[83.5-155.0] | **< 0.001** |
|  | 2018 | 61.8<br>[25.1-90.1] | 89.0<br>[29.0-164.0] | **< 0.001** |
|  | 2019 | 49.5<br>[20.1-66.1] | 135.0<br>[96.0-171.0] | **< 0.001** |
| Temperature °C (sending samples), median [IQR] | 2016 | 19.6<br>[18.1-21.2] | 17.8<br>[16.0-19.1] | **< 0.001** |
|  | 2017 | 21.9<br>[19.6-24.1] | 18.0<br>[16.0-19.5] | **< 0.001** |
|  | 2018 | 21.5<br>[19.3-23.8] | 18.3<br>[16.8-19.8] | **< 0.001** |
|  | 2019 | 20.6<br>[19.0-22.2] | 18.1<br>[16.6-19.2] | **< 0.001** |
| Temperature °C (receiving samples), median [IQR] | 2016 | 20.1<br>[18.8-21.4] | 19.6<br>[17.9-21.6] | **< 0.001** |
|  | 2017 | 22.1<br>[20.6-24.1] | 20.7<br>[19.5-22.9] | **< 0.001** |
|  | 2018 | 21.9<br>[20.0-23.9] | 20.4<br>[18.6-22.9] | **< 0.001** |
|  | 2019 | 20.7<br>[19.5-22.1] | 20.5<br>[18.9-22.6] | **< 0.001** |
| Delta Temperature °C (receiving x sending, median [IQR]) | 2016 | 0.54<br>[0.0-0.8] | 1.82<br>[0.0-3.7] | **< 0.001** |
|  | 2017 | 0.26<br>[0.0-0.1] | 2.72<br>[1.2-4.1] | **< 0.001** |
|  | 2018 | 0.37<br>[0.0-0.4] | 2.06<br>[0.9-4.1] | **< 0.001** |
|  | 2019 | 0.15<br>[0.0-0.2] | 2.35<br>[1.1-4.3] | **< 0.001** |

\*Setup A: contains Site 1 and 2, characterized by performing the collection and processing of samples in the same place. Setup B: contains Site 3, 4 and 5. It is characterized by performing the collection and processing of samples in distinct place; <sup>#</sup>Data were compared between the setups using the Mann-Whitney U test. Bold and italic font indicates statistical significance. <sup>\*</sup>The number of samples changes overtime for each Setup, as follow: 2016 (A = 190; B = 173), 2017 (A = 366; B = 258), 2018 (A = 505; B = 471), 2019 (A = 570; B = 333). <sup>£</sup>Data are shown as median and interquartile range (IQR).

Setup A had higher temperature at the time of specimen shipment compared to Setup B, but both Setups had similar temperatures at the time of specimen receipt/arrival. (**Figure 6A, 6B**). Although within the established standard, Setup A had a significantly higher temperature at the time of both shipment and receipt. Despite this, the temperature variation (Δ) was significantly greater in Setup B (**Figure 6C**). This can also be seen when analyzing the data by site (**Supplementary Figure 2**).

**Figure 6:**
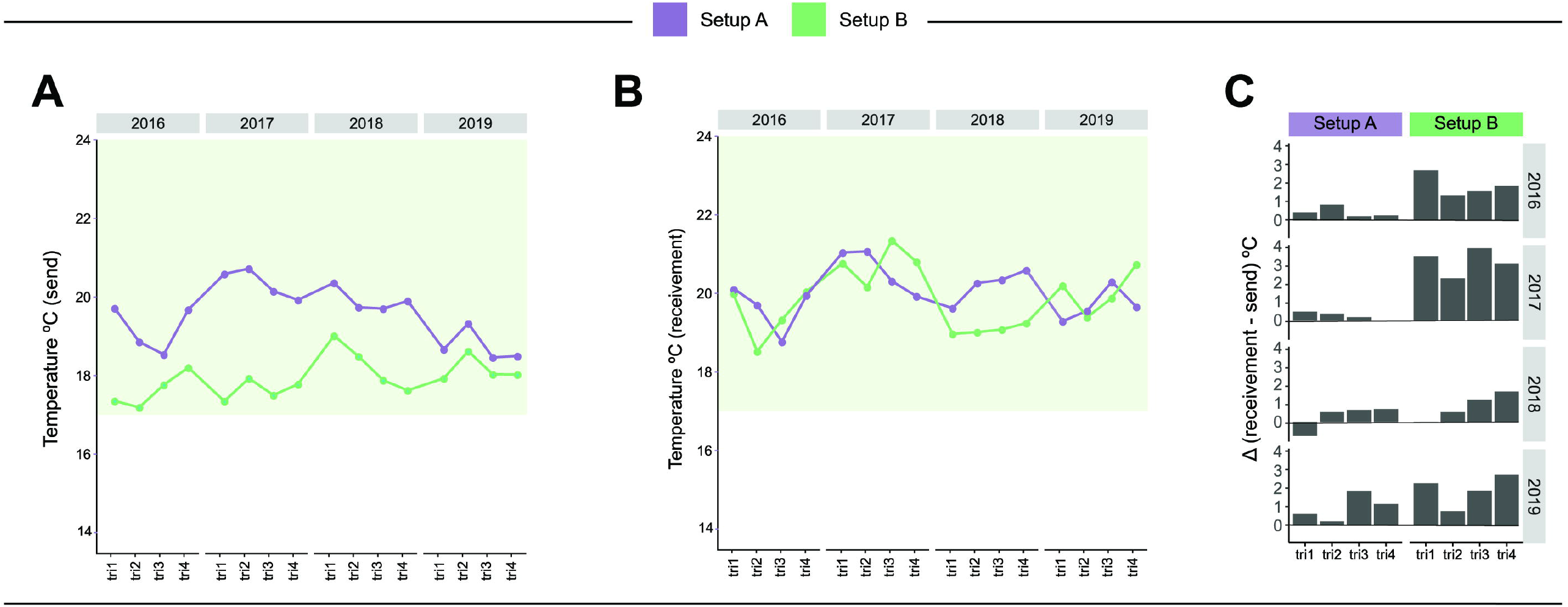
Dynamics of the temperature at the time of shipment and receipt of study samples, and the delta temperature variation over time in each Setup stratified by trimester and year during the study period. A) Average Temperature (°C) of sending samples calculated by trimester and year in each setup. B) Average Temperature (°C) of receiving samples calculated by trimester and year in each setup. C) The difference between receiving and sending temperature (Δ) was calculated for each trimester and year in each setup. Purple lines indicate Setup A and green lines indicate Setup B. The light green block indicates the limit accepted by the IGRA test manufacturer as acceptable for the storage and handling of the samples (17-27°C).

Over the years of implementation, the variation in time between sending and receiving samples tended to decrease until 2018, then there was an increase in Δ in 2019. The decrease in time variation was possibly because study and transport became more efficient (collection, sending and processing) with the implementation of the test on the sites. Although we observed this increase in 2019, it is noteworthy that there was an increase in the number of samples analyzed that year.

Finally, a correlation analysis was performed between the Δ (variation of receiving -sending) of time and temperature. In this analysis it was possible to note that the Δ°C was directly correlated with the Δ minutes (r = 0.37, p <0.001), and that this variation was particularly seen in Setup B (**Figure 7**).

**Figure 7:**
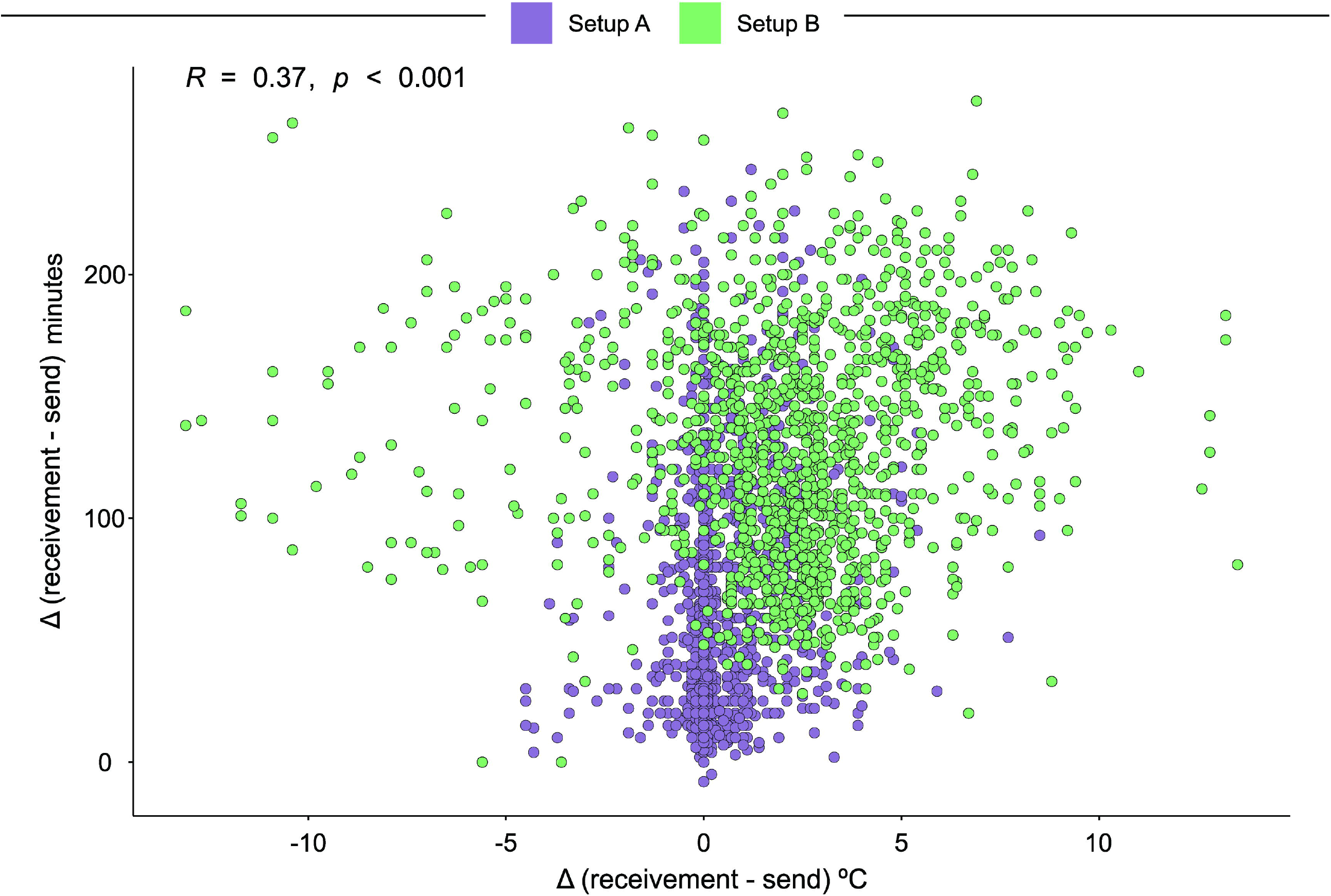
Correlation between delta temperature variation and delta time variation of study. Purple dots indicate Setup A and green dots indicate Setup B.

## 4 Discussion

In the IGRA implementation process, gaps were identified mainly in the pre-analytical phase of the QFT^®^-Plus assay. The training of laboratory technicians without experience in sample collection, transporting and processing, with ELISA assays, was straight-forward and the learning curve was quick, despite pre-analytical errors identified after the start of the study. Pre-analytical errors have important implications for the reproducibility and accuracy of IGRAs, indicating the need to standardize the pre-analytical steps, as shown previously (19). The sites standardized the preventive maintenance process of the equipment, to minimize problems. The delivery of the QFT^®^-Plus Tubes and ELISA kits were directed to one of the clinical sites, which then distributed them to the other laboratories, mitigating import and logistic issues. The steps of collection, transport and processing of the samples started to be monitored via Case Report Forms (CRFs). The lessons learned at this stage were important to create mechanisms for tracking non-conformities, It is noteworthy that the use of CRFs is indicated for monitoring non-conformities in clinical trials, however, they can also be used in routine laboratory conditions (20).

The sample collection, transport, and processing settings (Setup A and B) in the study influenced the results of the QFT^®^-Plus. Setup B had a higher percentage of indeterminate and positive results, probably due to the longer time between the collection and processing of the samples, in addition to other variables evaluated here (e.g., temperature of transport, ID of the tubes, and packaging of the samples). It is important to ensure that, in the analytical phase, the laboratory technicians involved in these steps are comfortable with performing QFT^®^-Plus, including simple tasks such as controlling the temperature of the shipping boxes, identifying non-conformities, minimizing errors, and identifying problems in the results generated. It is recommended to include in the QFT^®^-Plus implementation planning a period of adaptation and short retraining, directed at the critical stages of sample collection, transport and processing, with the aim of gradually minimizing or eliminating non-conformities and pre-analytical errors. This learning can assist in the quality control of test results and performance, since the reproducibility of IGRA can be influenced by these factors (21).

Biological samples can have different performance or results with respect to the quantity of analytes used and based on this, we must seek the identification of points that cause variation of test results. The laboratory routine can have different components that can cause variability in results, such as different processing rates, variation in processing rates throughout the month, and variation of factors related to the environment (such as temperature) throughout the year (22).

The reproducibility of the test results depends directly on the training of the team, since variation in the “operator” for the collection, homogenization and performance of the test can impact the result. Therefore, the standardization of quality control in the clinical and laboratory spheres is essential so that there are no significant effects on the result. In addition, structural variations between different laboratories can influence the proportion of indeterminate results (21, 23).

With regard to immunological molecules that can be released and consumed quickly (in vitro), delays in the start of the incubation of study samples can interfere in the quantification of IFN-γ levels, leading to a decrease of up to 0.24 IU/mL after 6 hours (19, 21). Different incubation times, without a specific pattern, can possibly influence test results. Therefore, it is important to follow the manufacturer’s guidelines (21, 23). The required quantity of final volume of biological sample must always be used, and it is not possible to use volumes smaller or larger than that recommended for the test. Due to the amount of “biological stimulus” available per tube, the amount of IFN-γ released at the end of the test may be affected by variation in the sample volume used (21, 24, 25).

The expansion of the use of IGRAs, such as QFT^®^-Plus, must be well planned, with negotiations with the manufacturer regarding the logistics of delivery of the kits in areas with difficult access. In addition, the sample collection, transport, and processing settings must be evaluated, with the aim of mitigating errors that may interfere with the test results. Training of laboratory technicians is extremely important, and regular training develops a sense of responsibility towards reporting non-conformities and maintaining data quality after initial implementation. Finally, we believe that some of the problems identified in this study can assist laboratories wishing to implement IGRAs, in addition to manufacturers of IGRAs providing effective technical support. These findings may facilitate the implementation of IGRA testing in countries with a high TB burden, such as Brazil.

## Data Availability

The original contributions presented in the study are included in the article/supplementary material, further inquiries can be directed to the corresponding author/s.

## 6 Author Contributions

AGC and MCS established the initial conception and wrote this manuscript. AGC, BKSC, MA-P, HNSI, RS-G, ABS, AG-S, AMSA, ECS, AB, MSR, ASRM, JG and, MCF collected, analyzed, and reviewed the data. AGC, BKSC, MA-P, MMT analyzed data and designed the illustrations and tables. MCF, BD, SC, ALK, VCR, TRS, BBA, MCS supervised the project development, interpreted the data, and reviewed this manuscript. All authors read, discussed the general outline of the article together and approved the final manuscript.

## 8 Acknowledgments

The authors thank the study participants. We also thank the teams of clinical and laboratory platforms of RePORT-Brazil. A special thanks to Elze Leite (FIOCRUZ, Salvador, Brazil), Eduardo Gama (FIOCRUZ, Rio de Janeiro, Brazil), Elcimar Junior (FMT-HVD, Manaus, Brazil), and Hilary Vansell (VUMC, Nashville, USA) for administrative and logistical support.

## 10 Contributors: Participating Investigators in RePORT Brazil Consortium

The RePORT Brazil Consortium consists of 12 partner institutions from Brazil represented by the following members: Amanda Araújo da Costa (A.A. Costa), Instituto de Pesquisa Clínica Carlos Borborema, Fundação de Medicina Tropical Doutor Heitor Vieira Dourado, Manaus, Brazil; André Luiz Bezerra (A.L. Bezerra), Faculdade de Medicina, Universidade Federal do Rio de Janeiro, Rio de Janeiro, Brazil; Anna Cristina Calçada Carvalho (A.C.C Carvalho), Faculdade de Medicina, Universidade Federal do Rio de Janeiro, Rio de Janeiro, Brazil, Laboratório de Inovações em Terapias, Ensino e Bioprodutos, Fundação Oswaldo Cruz, Rio de Janeiro, Brazil; Anna Karla Silveira (A.K. Silveira), Faculdade de Medicina, Universidade Federal do Rio de Janeiro, Rio de Janeiro, Brazil; Betânia M. F. Nogueira (B.M.F. Nogueira), Instituto Brasileiro para Investigação da Tuberculose, Fundação José Silveira, Salvador, Brazil, Multinational Organization Network Sponsoring Translational and Epidemiological Research (MONSTER) Initiative, Salvador, Brazil, Faculdade de Medicina, Universidade Federal da Bahia, Salvador, Brazil; Bruna da Costa Oliveira Lima (B.C.O. Lima), Instituto de Pesquisa Clínica Carlos Borborema, Fundação de Medicina Tropical Doutor Heitor Vieira Dourado, Manaus, Brazil; Bruna Pires de Loiola (B.P. Loiola), Instituto de Pesquisa Clínica Carlos Borborema, Fundação de Medicina Tropical Doutor Heitor Vieira Dourado, Manaus, Brazil; Carolina Arana Schmaltz Stanis (C.A. Schmaltz), Instituto Nacional de Infectologia Evandro Chagas, Fundação Oswaldo Cruz, Rio de Janeiro, Brazil; Eline Naiane de Freitas Medeiros (E.N.F. Medeiros), Instituto de Pesquisa Clínica Carlos Borborema, Fundação de Medicina Tropical Doutor Heitor Vieira Dourado, Manaus, Brazil; Francine Peixoto Ignácio (F.P. Ignácio), Instituto Nacional de Infectologia Evandro Chagas, Fundação Oswaldo Cruz, Rio de Janeiro, Brazil; Hayna Malta Santos (H.M. Santos), Faculdade de Medicina, Universidade Federal da Bahia, Salvador, Brazil, Laboratório de Inflamação e Biomarcadores, Instituto Gonçalo Moniz, Fundação Oswaldo Cruz, Salvador, Brazil; Jéssica Rebouças Silva (J.R. Silva), Faculdade de Medicina, Universidade Federal da Bahia, Salvador, Brazil, Laboratório de Inflamação e Biomarcadores, Instituto Gonçalo Moniz, Fundação Oswaldo Cruz, Salvador, Brazil; João Marine Neto (J.M. Neto), Secretaria Municipal de Saúde do Rio de Janeiro - SMS-RJ - Rio de Janeiro, Brazil, Hospital Federal do Andaraí - Ministério da Saúde, Brazil; Leandro Sousa Garcia (L.S. Garcia), Instituto de Pesquisa Clínica Carlos Borborema, Fundação de Medicina Tropical Doutor Heitor Vieira Dourado, Manaus, Brazil; Maria Luciana Silva-Freitas (M.L. Silva-Freitas), Laboratório Interdisciplinar de Pesquisas Médicas, Instituto Oswaldo Cruz, FIOCRUZ, Rio de Janeiro, RJ; Mayla Gabriele Miranda de Melo (M.G.M. Melo), Faculdade de Medicina, Universidade Federal do Rio de Janeiro, Rio de Janeiro, Brazil; Rosa Maria Placido-Pereira (R.S. Placido-Pereira), Laboratório Interdisciplinar de Pesquisas Médicas, Instituto Oswaldo Cruz, FIOCRUZ, Rio de Janeiro, RJ; Samyra Almeida-Da-Silveira (S. Almeida-Da-Silveira), Laboratório Interdisciplinar de Pesquisas Médicas, Instituto Oswaldo Cruz, FIOCRUZ, Rio de Janeiro, RJ; Vanessa de Souza Nascimento (V.S. Nascimento), Instituto Brasileiro para Investigação da Tuberculose, Fundação José Silveira, Salvador, Brazil; 5. Multinational Organization Network Sponsoring Translational and Epidemiological Research (MONSTER) Initiative, Salvador, Brazil, Bahiana School of Medicine and Public Health, Bahia Foundation for the Development of Sciences, Salvador, Brazil.

